# The night-time sleep and autonomic activity of male and female professional road cyclists competing in the Tour de France and Tour de France Femmes

**DOI:** 10.1101/2023.07.06.23292280

**Authors:** Charli Sargent, Summer Jasinski, Emily R. Capodilupo, Jeremy Powers, Dean J. Miller, Gregory D. Roach

**Author notes:** Corresponding Author:* Charli Sargent –.

## Abstract

**Objective:** The objective of this study was to examine the capacity of male and female professional cyclists to recover between daily race stages while competing in the 109^th^ edition of the Tour de France (2022) and the 1^st^ edition of the Tour de France Femmes (2022), respectively.

**Methods:** The 17 participating cyclists were 8 males from a single team (aged 28.0 ± 2.5 years [mean ± 95% confidence interval]) and 9 females from two separate teams (aged 26.7 ± 3.1 years). Cyclists wore a fitness tracker (WHOOP 4.0) to capture recovery metrics primarily related to night-time sleep and autonomic activity. Data were collected for the entirety of the events and for 7 days of baseline before the events. The primary analyses tested for a main effect of ‘stage type’ – i.e., rest, flat, hilly, mountain or time trial for males and flat, hilly or mountain for females – on the various recovery metrics.

**Results:** During baseline, total sleep time at night was 7.2 ± 0.3 h for male cyclists and 7.7 ± 0.3 h for female cyclists, sleep efficiency (i.e., total sleep time as a percentage of time in bed) was 87.0 ± 4.4 % for males and 88.8 ± 2.6 % for females, resting heart rate was 41.8 ± 4.5 beats·min^-1^ for males and 45.8 ± 4.9 beats·min^-1^ for females, and heart rate variability during sleep was 108.7 ± 17.0 ms for males and 119.8 ± 26.4 ms for females. During their respective events, total sleep time at night was 7.2 ± 0.1 h for males and 7.5 ± 0.3 h for females, sleep efficiency was 86.4 ± 1.2 % for males and 89.6 ± 1.2 % for females, resting heart rate was 44.5 ± 1.2 beats·min^-1^ for males and 50.2 ± 2.0 beats·min^-1^ for females, and heart rate variability during sleep was 99.1 ± 4.2 ms for males and 114.3 ± 11.2 ms for females. For male cyclists, there was a main effect of ‘stage type’ on recovery, such that heart rate variability during sleep was lowest after mountain stages. For female cyclists, there was also a main effect of ‘stage type’ on recovery, such that the percentage of light sleep in a sleep period (i.e., lower-quality sleep) was highest after mountain stages.

**Conclusions:** Some aspects of recovery were compromised in cyclists after the most demanding days of racing, i.e., mountain stages. Overall however, the cyclists obtained a reasonable amount of good-quality sleep while competing in these highly demanding endurance events. This study demonstrates that it is now feasible to assess recovery metrics in professional athletes during multiple-day endurance events using validated fitness trackers.

## 1 Introduction

In professional men’s cycling, the most physically and mentally demanding events are the Grand Tours – i.e., Tour de France, Giro d’Italia and Vuelta a España [1]. The Grand Tours are held over three consecutive weeks of racing with daily stages covering a variety of terrain – flat, hilly and mountain. Typically, the Tour de France imposes the highest total exercise loads on its participants due to the duration and length of the daily stages – the longest by duration last up to 5 hours and the longest by length cover up to 300 km [2, 3]. In recent decades, competitors in the Tour de France have covered an average distance of 3,650 ± 208 km, with the winners taking an average of 92 ± 6 hours to complete the race [1]. There are no three-week tours in women’s professional cycling, but the most prestigious events, and potentially the most demanding, are the one-week tours that are ‘sister’ events to the men’s Grand Tours, known as Tour de France Femmes, Giro Donne and Vuelta Femenina.

The physiological demands of the Grand Tours are well above those that can be undertaken by most endurance-trained athletes [2]. To this end, the requirements of the main types of stages during Grand Tours have been well described in terms of both the volume and intensity of exercise [1–7]. For example, exercise duration is shortest for time-trial stages and longest for flat and mountain stages; exercise intensity is low to moderate during flat stages, moderate to high during mountain stages, and high during time-trial stages; and physiological load is greatest during mountain stages compared to flat and time-trial stages [1, 4]. Understanding the physiological demands imposed on cyclists during Grand Tours is useful in terms of the design of training strategies to optimise potential performance [4], but this information does not provide insight into the capacity of cyclists to physiologically recover between stages.

Sleep is considered one of the major forms of recovery from exercise [8], but it can be disrupted following single bouts of ultra-endurance exercise [9]. Obtaining sufficient sleep may be particularly challenging during Grand Tours because cyclists must complete extreme bouts of endurance exercise each day with little time to recover before the next stage of racing. Specifically, road cyclists report needing 8.2 hours of sleep per night to feel fully rested [10], but during one-week multi-stage races, they obtain an average of only ∼7 hours of sleep per night [11, 12]. Furthermore, in simulated ‘Grand Tours’, average sleep duration declines over consecutive weeks of racing from 7.4 hours to 7.0 hours [13]. In addition, indicators of recovery based on autonomic activity, i.e., resting heart rate and heart rate variability, also decline over consecutive weeks of simulated racing [14].

For cyclists competing in Grand Tours, insufficient recovery between stages because of poor sleep could affect performance during subsequent stages of the race. The physiological demands placed on cyclists during Grand Tours have been quantified, but the impact of these demands on daily recovery in terms of sleep and autonomic activity have yet to be determined. The emergence of wearable technologies now makes it possible to monitor sleep and autonomic activity non-invasively during competition [15]. In the present study, data were collected using one such wearable technology, i.e., WHOOP 4.0 (Whoop, Inc., Boston, MA, USA), with professional male and female cyclists competing in the 2022 editions of the Tour de France and Tour de France Femmes, respectively. The aims of the study were to (i) quantify the impact of race days on sleep and autonomic activity in professional male and female cyclists; (ii) examine whether the characteristics of a race stage – i.e., flat, hilly, mountain, time trial – differentially affects sleep and autonomic activity in professional male and female cyclists; and (iii) determine whether there is a cumulative effect of racing over consecutive weeks on sleep and autonomic activity in professional male cyclists.

## 2 Procedure

### 2.1 Participants

Seventeen professional road cyclists from teams competing in the 2022 editions of the Tour de France and Tour de France Femmes provided informed consent for their data to be included in the present study. The male riders (*n* = 8; age: 28.0 ± 2.5 years; mass: 70.3 ± 4.6 kg; height: 181.6 ± 4.9 cm; mean ± 95% confidence interval) were members of the same team. The female riders (*n* = 9; age: 26.7 ± 3.1 years; mass: 58.0 ± 4.4 kg; height: 169.8 ± 5.6 cm; mean ± 95% confidence interval) were members of two teams (team 1: *n* = 4; team 2: *n* = 5). The procedure for data collection complied with the Declaration of Helsinki and was approved by CQUniversity’s Human Research Ethics Committee.

### 2.2 Race Schedule

#### 2.2.1 Males

In 2022, the Tour de France included 21 stages of racing from the 1^st^ of July to the 24^th^ of July (Table 1). The race consisted of two individual time trials (stages 1, 20), six flat stages (2, 3, 13, 15, 19, 21), seven hilly stages (4–6, 8, 10, 14, 16), six mountain stages (7, 9, 11, 12, 17, 18), two rest days, and one transfer day – with a total distance of 3,581 km and a total elevation gain of 47,026 m.

**Table 1.**
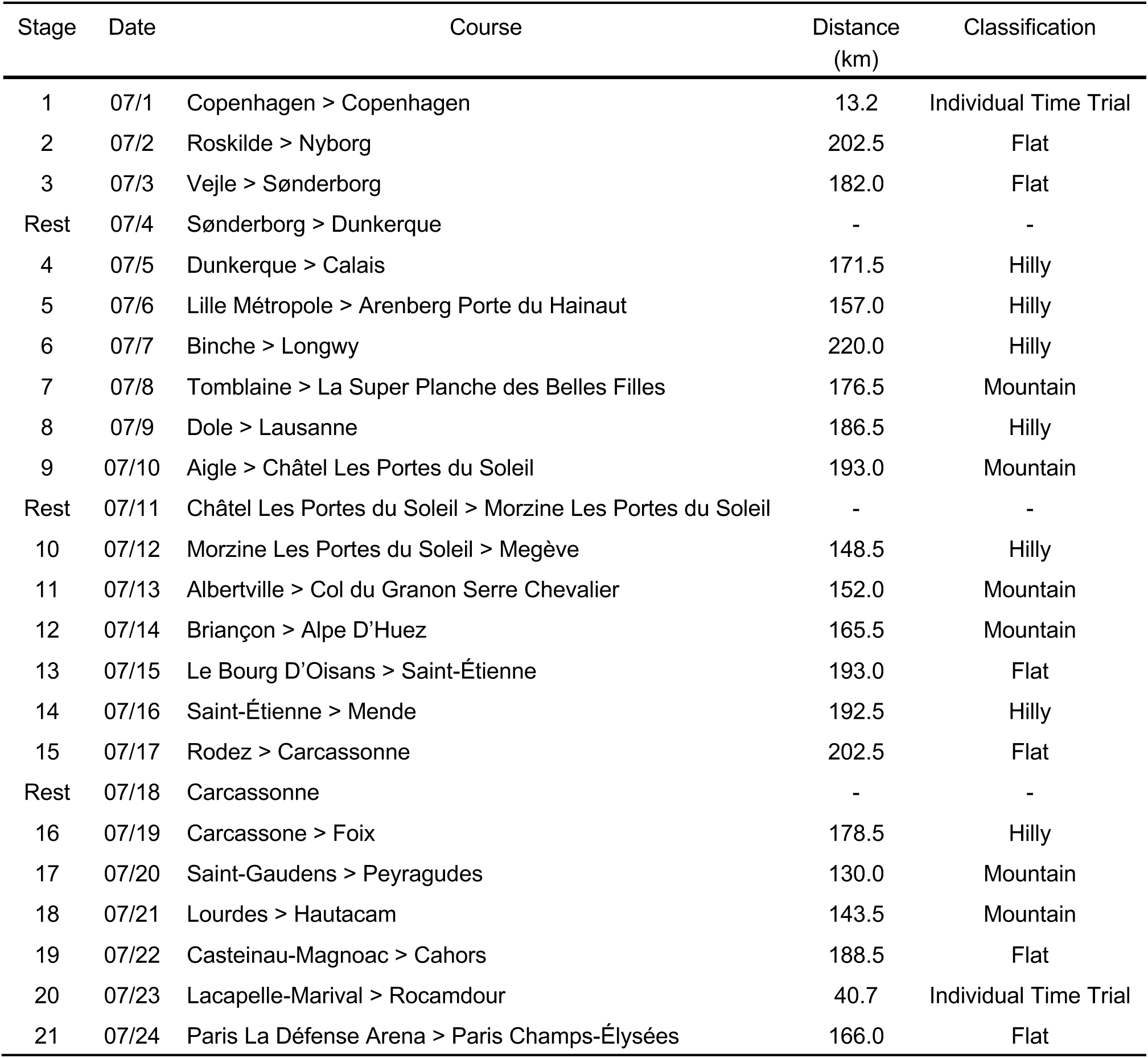
2022 Tour de France Route and Classification.

| Stage | Date | Course | Distance (km) | Classification |
| --- | --- | --- | --- | --- |
| 1 | 07/1 | Copenhagen > Copenhagen | 13.2 | Individual Time Trial |
| 2 | 07/2 | Roskilde > Nyborg | 202.5 | Flat |
| 3 | 07/3 | Vejle > Sønderborg | 182.0 | Flat |
| Rest | 07/4 | Sønderborg > Dunkerque | - | - |
| 4 | 07/5 | Dunkerque > Calais | 171.5 | Hilly |
| 5 | 07/6 | Lille Métropole > Arenberg Porte du Hainaut | 157.0 | Hilly |
| 6 | 07/7 | Binche > Longwy | 220.0 | Hilly |
| 7 | 07/8 | Tomblaine > La Super Planche des Belles Filles | 176.5 | Mountain |
| 8 | 07/9 | Dole > Lausanne | 186.5 | Hilly |
| 9 | 07/10 | Aigle > Châtel Les Portes du Soleil | 193.0 | Mountain |
| Rest | 07/11 | Châtel Les Portes du Soleil > Morzine Les Portes du Soleil | - | - |
| 10 | 07/12 | Morzine Les Portes du Soleil > Megève | 148.5 | Hilly |
| 11 | 07/13 | Albertville > Col du Granon Serre Chevalier | 152.0 | Mountain |
| 12 | 07/14 | Briançon > Alpe D'Huez | 165.5 | Mountain |
| 13 | 07/15 | Le Bourg D'Oisans > Saint-Étienne | 193.0 | Flat |
| 14 | 07/16 | Saint-Étienne > Mende | 192.5 | Hilly |
| 15 | 07/17 | Rodez > Carcassonne | 202.5 | Flat |
| Rest | 07/18 | Carcassonne | - | - |
| 16 | 07/19 | Carcassonne > Foix | 178.5 | Hilly |
| 17 | 07/20 | Saint-Gaudens > Peyragudes | 130.0 | Mountain |
| 18 | 07/21 | Lourdes > Hautacam | 143.5 | Mountain |
| 19 | 07/22 | Casteinau-Magnoac > Cahors | 188.5 | Flat |
| 20 | 07/23 | Lacapelle-Marival > Rocamadour | 40.7 | Individual Time Trial |
| 21 | 07/24 | Paris La Défense Arena > Paris Champs-Élysées | 166.0 | Flat |

#### 2.2.2 Females

In 2022, the Tour de France Femmes included eight stages of racing from the 24^th^ of July to the 31^st^ of July (Table 2). The race did not have any time trials or rest days, but it consisted of three flat stages (1, 2, 5), three hilly stages (3, 4, 6), and two mountain stages (7, 8) – with a total distance of 1,033 km and a total elevation gain of 12,707 m.

**Table 2.** 2022 Tour de France Femmes Route and Classification.

| Stage | Date | Course | Distance (km) | Classification |
| --- | --- | --- | --- | --- |
| 1 | 07/24 | Tour Eiffel > Paris Champs-Élysées | 81.6 | Flat |
| 2 | 07/25 | Meaux > Provins | 136.4 | Flat |
| 3 | 07/26 | Reims > Épernay | 133.5 | Hilly |
| 4 | 07/27 | Troyes > Bar-Sur-Aube | 126.8 | Hilly |
| 5 | 07/28 | Bar-Le-Duc > Saint-Dié-Des-Vosges | 175.6 | Flat |
| 6 | 07/29 | Saint-Dié-Des-Vosges > Rosheim | 129.2 | Hilly |
| 7 | 07/30 | Sélestat > Le Markstein | 127.1 | Mountain |
| 8 | 07/31 | Lure > La Super Planche Des Belles Filles | 123.3 | Mountain |

### 2.3 Sleep and Cardiac Activity

The participants’ sleep and cardiac activity were monitored using a fitness tracker worn on the wrist (WHOOP 4.0, CB Rank, Boston, MA, USA). For each night-time sleep opportunity, the following sleep variables were obtained: sleep onset time, sleep offset time, time in bed (time between sleep onset and sleep offset), total sleep time, sleep efficiency (total sleep time / time in bed x 100), light sleep (expressed as a percentage of time in bed), slow wave sleep (expressed as a percentage of time in bed), and rapid eye movement (REM) sleep (expressed as a percentage of time in bed). Resting heart rate (beats·min^-1^) was calculated as the mean heart rate of non-wake periods of the primary sleep episode. Heart rate variability (rMSSD, ms) was calculated using the root-mean-square of successive differences between heartbeats formula across the non-wake periods of the primary sleep episode. In all analyses, the sleep and cardiac activity associated with a particular stage are the data collected on the night immediately following that stage.

Previous generations of the WHOOP strap (WHOOP 2.0 and 3.0) have been validated against the gold standard for monitoring sleep (i.e., polysomnography) and cardiac activity (i.e., electrocardiogram) [15–17]. Compared with polysomnography, WHOOP 3.0’s agreement (and Cohen’s kappa) is 86% (κ = 0.44) for two-state categorisation of sleep periods (as sleep or wake) and 60% (κ = 0.44) for multistate categorisation of sleep periods (as a specific sleep stage or wake). Compared with electrocardiogram, WHOOP 3.0 underestimates resting heart rate by an average of 0.3 beats·min^-1^ and underestimates heart rate variability (rMSSD) by an average of 4.5 ms. For both heart rate and heart rate variability, the intraclass correlation between WHOOP 3.0 and electrocardiogram is 0.99, i.e., excellent.

In addition to the sleep and cardiac variables described above, WHOOP ‘daily strain’ was calculated for each stage of the race. WHOOP daily strain measures ‘total cardiovascular load’ on a proprietary, non-linear scale of 0 to 21 and is categorised into the following bands: light strain (0-9), moderate strain (10-13), high strain (14-17), and overreaching (18-21) [18].

### 2.4 Statistical Analyses

The aims of the study were addressed by conducting a series of linear mixed effects models using the variance components covariance structure and restricted maximum likelihood estimation. Statistical analyses were performed using SPSS (version 28.0.0.0; IBM; Armonk, NY, USA). Results are reported as mean (± 95% confidence intervals) and were considered significant at *p* < .05. Separate linear mixed effects models were conducted for males and females.

Changes in sleep and cardiac variables during each race were examined by entering ‘day of race’ as a fixed effect into the model and participant as a random effect. The fixed effect in the models (i.e., ‘day of race’) included baseline (i.e., average value for each participant calculated from Monday to Sunday at least one week prior to the first day of the race), day of race (21 race days, 1 transfer day and 2 rest days) and post-race (i.e., average value for each participant calculated from post-race night 2 to post-night 9). Where a main effect of ‘day of race’ was observed, pairwise comparisons with a Bonferroni correction were performed with ‘baseline’ as the reference category.

The impact of ‘stage classification’ on the dependent variables was examined using linear mixed effects models. In the models, ‘participant’ was entered as a random effect and ‘stage classification’ was entered as a fixed effect. Stage classifications were based on the categorisations published by the race organisers. For the males, the fixed effect ‘stage classification’ included rest (transfer day, rest day 1, rest day 2), flat (stages 2, 3, 13, 15, 19), hilly (stages 4–6, 8, 10, 14, 16), mountain (stages 7, 9, 11, 12, 17, 18) and individual time trial (stages 1, 20). For the females, the fixed effect ‘stage classification’ included flat (stages 1, 5), hilly (stages 2, 3, 6), and mountain (stages 4, 7). The variables obtained on the night of the final stage for the males (stage 21) and the females (stage 8) were excluded from the analyses. Where a main effect of ‘stage classification’ was observed, all pairwise comparisons were performed with a Bonferroni correction applied.

For the males, additional linear mixed effects models were performed to examine the impact of ‘week of race’ on the dependent variables. In the models, ‘participant’ was entered as a random effect and ‘week of race’ was entered as a fixed effect. The fixed effect ‘week of race’ included baseline (calculated as described above), week 1 (stages 1 to 9 but excluding the transfer day), week 2 (stages 10 to 15 but excluding the first rest day) and week 3 (stages 16 to 20 but excluding the second rest day). The variables obtained on the night of the final stage (stage 21) were excluded from the analyses. Where a main effect of ‘week of race’ was observed, all pairwise comparisons were performed with a Bonferroni correction applied.

## 3 Results

### 3.1 Race Completion and Compliance

Of the eight male riders who participated in data collection, six completed all 21 stages of the race and two riders withdrew from the race. For the two riders that withdrew from the race, only nights following successful completion of a stage were included in the analyses and all subsequent nights after racing were excluded from the analyses. Of the 56 nights available to calculate a baseline average, 55 nights were collected (98%); of the 166 nights available following successful completion of a stage of the race, 159 were collected (96%); and of the 56 nights available to calculate a post-race average, 48 were collected (86%).

Of the nine female riders who participated in data collection, eight completed all eight stages of the race, and one rider withdrew from the race. For the rider that withdrew from the race, only nights following successful completion of a stage were included in the analyses and all subsequent nights after racing were excluded from the analyses. Of the 63 nights available to calculate a baseline average, 63 were collected (100%); of the 70 nights available following successful completion of a stage of the race, 66 were collected (94%); and of the 63 nights available to calculate a post-race average, 61 were collected (97%).

### 3.2 Male Cyclists

#### 3.2.1 Baseline

For each participant, data were collected at least one week prior to the start of the first stage of the race and included five weekdays (Monday-Friday) and two weekend days (Saturday, Sunday). On average, the participants fell asleep at 22:03 h ± 40.8 min, woke at 06:21 h ± 43.2 min, spent 8.3 ± 0.2 h in bed, and obtained 7.2 ± 0.3 h of sleep with an efficiency of 87.0 ± 4.4 %. The percentage of time in bed spent in each stage of sleep was 45.1 ± 5.5 % for light sleep, 18.5 ± 2.5 % for slow wave sleep and 23.4 ± 4.3 % for REM sleep. In comparison, for male/female non-athletes of a similar age (25 years), the percentage of time in bed spent in each stage of sleep is 54.8 % for light sleep, 16.3 % for slow wave sleep and 20.7 % for REM sleep [19]. The participants’ average resting heart rate was 41.8 ± 4.5 beats·min^-1^ and average heart rate variability was 108.7 ± 17.0 ms. In comparison, male/female non-athletes of a similar age (20–29 years) have resting heart rate of 54.2 beats·min^-1^ and heart rate variability of 82.9 ms [20].

#### 3.2.2 Day of Race (Fig. 1–4)

On average during the race, the participants fell asleep at 22:18 h ± 7.8 min, woke at 06:36 h ± 5.4 min, spent 8.4 ± 0.2 h in bed, and obtained 7.2 ± 0.1 h of sleep with an efficiency of 86.4 ± 1.2 %. The percentage of time in bed spent in each stage of sleep was 49.5 ± 1.7 % for light sleep, 17.3 ± 0.7 % for slow wave sleep and 19.6 ± 1.4 % for REM sleep. The participants’ average resting heart rate was 44.5 ± 1.2 beats·min^-1^ and average heart rate variability was 99.1 ± 4.2 ms. There was a main effect of ‘day of race’ on sleep onset time, sleep offset time, time in bed, and total sleep time (Table 3). Compared with baseline, participants fell asleep later on the night of the final stage (+3.2 h; *p* < .001), spent less time in bed on the night of the final stage (-2.7 h; *p* < .001), and obtained less sleep on the night of the final stage (-2.1 h; *p* < .008). There was a main effect of ‘day of race’ on sleep offset time but none of the pairwise comparisons were significant. There was a main effect of ‘day of race’ on daily strain. Compared with baseline, daily strain was higher on all days except for stage 1 and stage 20, the three rest days, and post-race. Daily strain was not different from baseline for stage 1, rest days 1, 2 and 3, stage 20 and post-race. There was no main effect of ‘day of race’ on resting heart rate or heart rate variability.

**Fig. 1.**
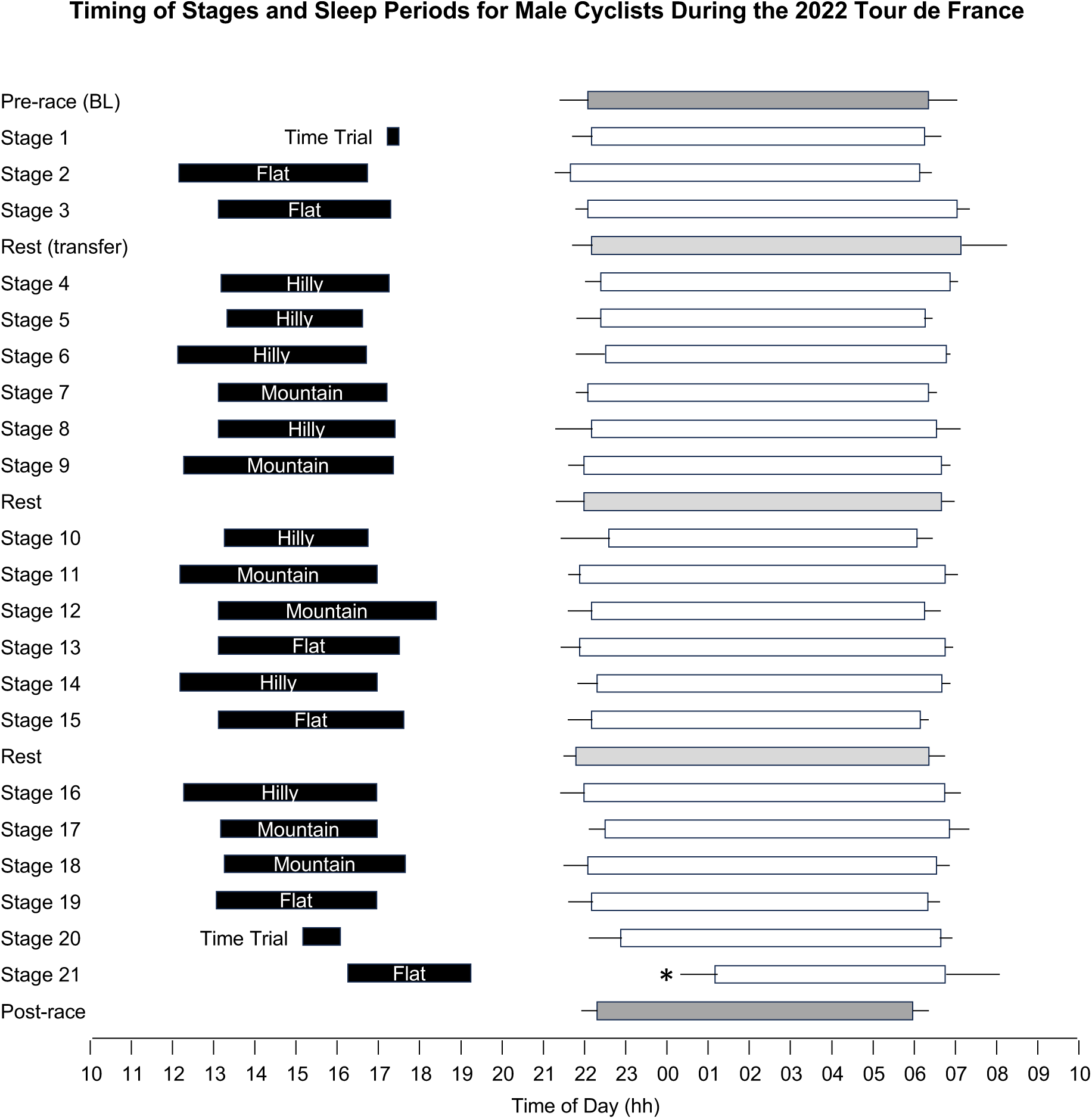
Timing of stages and sleep periods for male cyclists during the 2022 Tour de France. Each line represents a 24-hour day from 10:00 AM to 10:00 AM. Black bars indicate the start/end times and duration of stages, based on the cyclists who participated in the study. The start/end times and duration of sleep periods for participating cyclists are represented by white bars for sleep after race stages, light grey bars for sleep after rest days and dark grey bars for sleep in the 14 days before and after competition. Data are mean and 95% confidence intervals. * indicates a significant difference from baseline (*p* < .05).

**Fig. 2.**
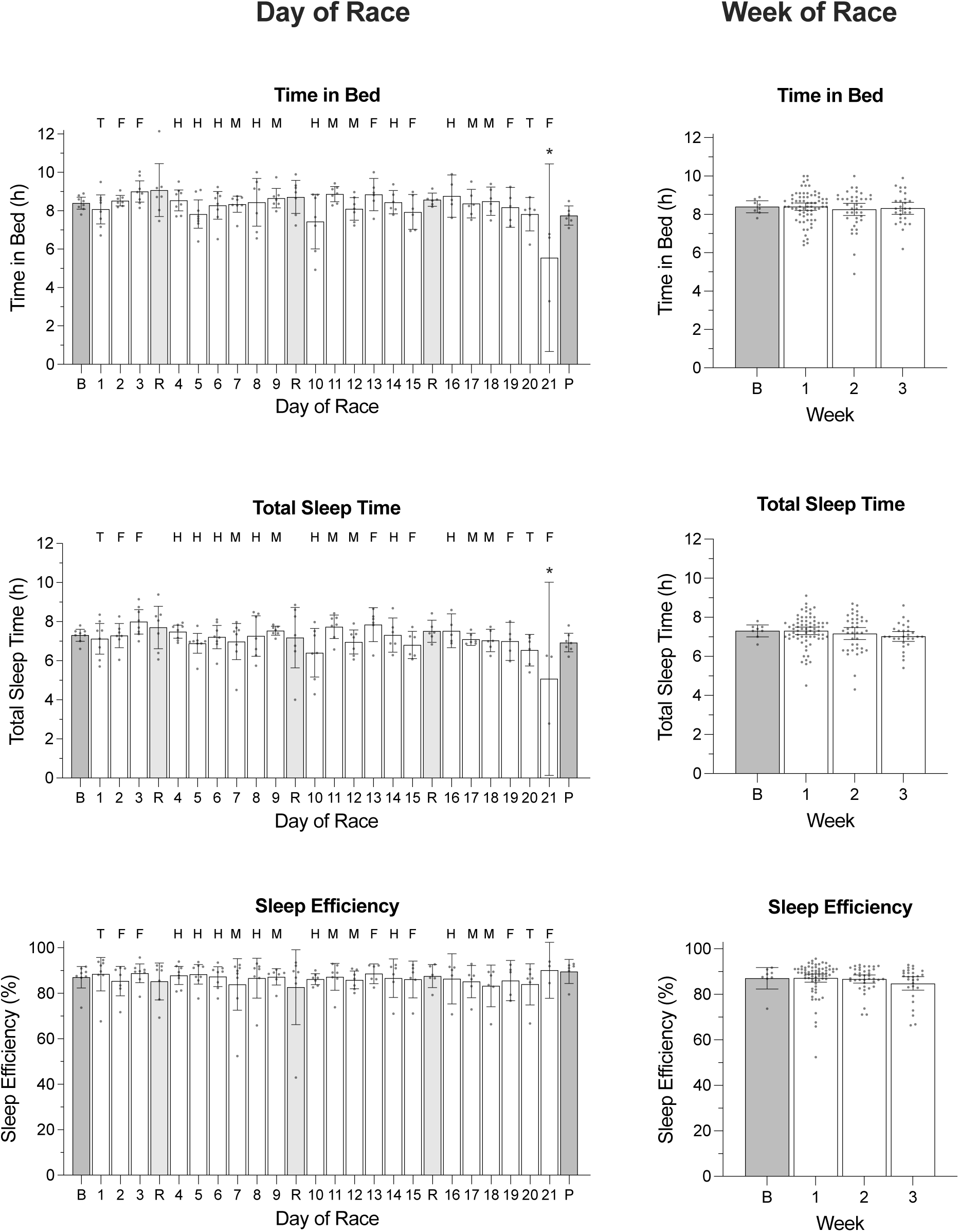
Time in bed, total sleep time, and sleep efficiency of male cyclists during the Tour de France plotted as a function of day of race (left panel) and week of race (right panel). ‘B’ represents the baseline average calculated at least one week prior to the first day of the race; ‘P’ indicates the post-race average calculated from post-race night 2 to post-night 9; ‘R’ indicates rest days; ‘T’ indicates individual time trial; ‘F’ indicates flat stages; ‘H’ indicates hilly stages; ‘M’ indicates mountain stages. Data are presented as mean (bars) and 95% confidence intervals (error bars). Closed circles represent individual cyclists. In the left panel, * indicates a significant difference from baseline (*p* < .05).

**Fig. 3.**
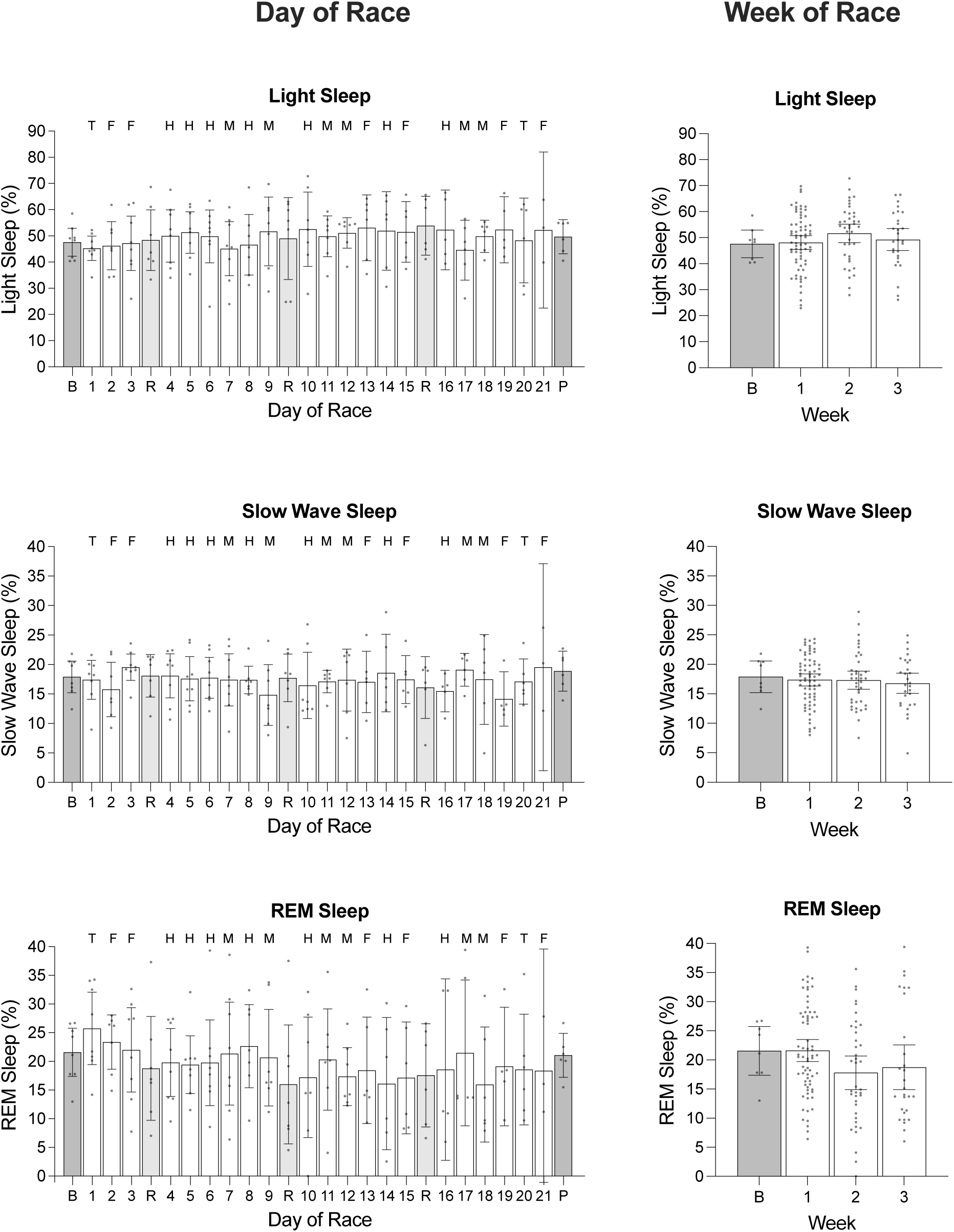
The percentage of light sleep, slow wave sleep and REM sleep obtained by male cyclists during the Tour de France plotted as a function of day of race (left panel) and week of race (right panel). ‘B’ represents the baseline average calculated at least one week prior to the first day of the race; ‘P’ indicates the post-race average calculated from post-race night 2 to post-night 9; ‘R’ indicates rest days; ‘T’ indicates individual time trial; ‘F’ indicates flat stages; ‘H’ indicates hilly stages; ‘M’ indicates mountain stages. Data are presented as mean (bars) and 95% confidence intervals (error bars). Closed circles represent individual cyclists.

**Fig. 4.**
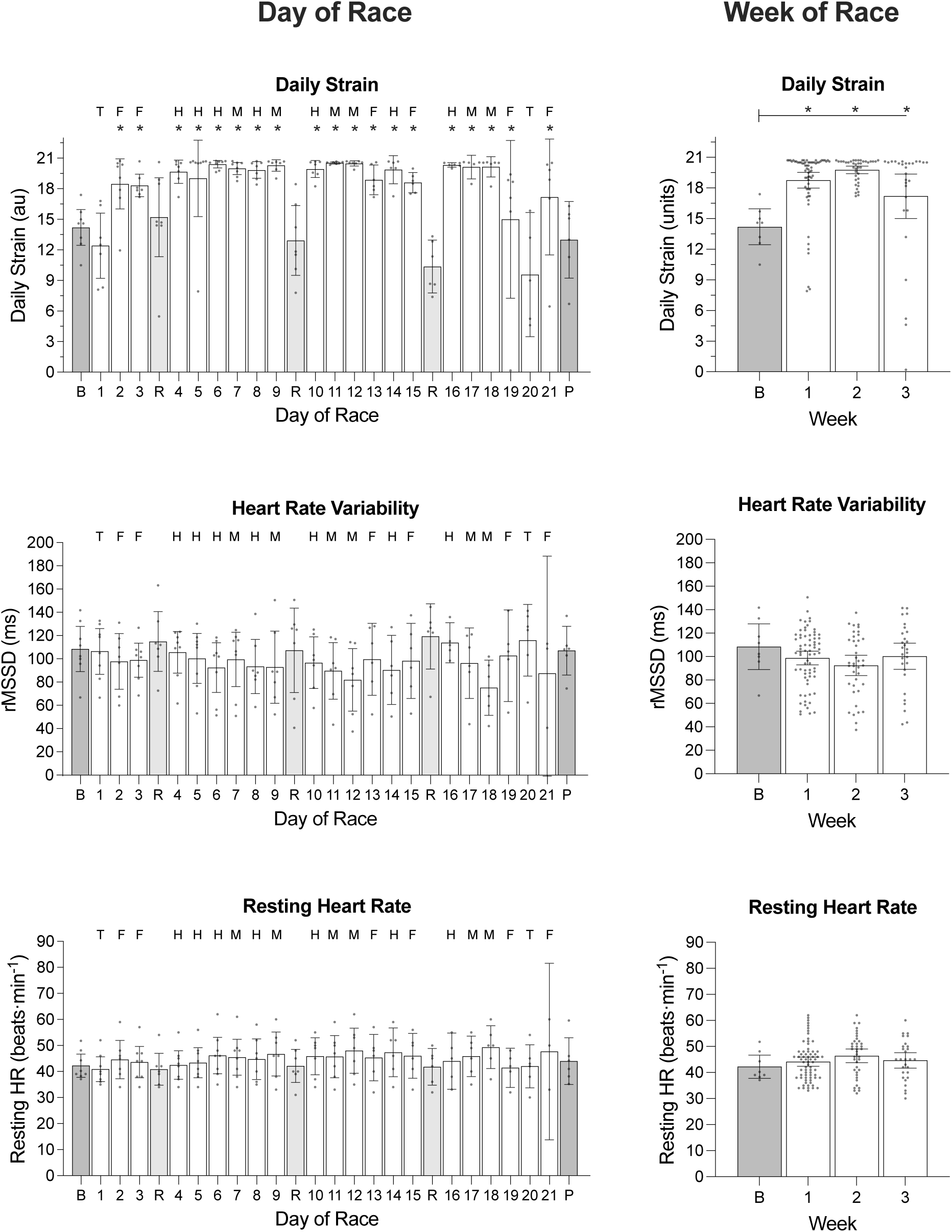
Daily strain, heart rate variability and resting heart rate of male cyclists during the Tour de France plotted as a function of day of race (left panel) and week of race (right panel). ‘B’ represents the baseline average calculated at least one week prior to the first day of the race; ‘P’ indicates the post-race average calculated from post-race night 2 to post-night 9; ‘R’ indicates rest days; ‘T’ indicates individual time trial; ‘F’ indicates flat stages; ‘H’ indicates hilly stages; ‘M’ indicates mountain stages. Data are presented as mean (bars) and 95% confidence intervals (error bars). Closed circles represent individual cyclists. In the left and right panels, * indicates a significant difference from baseline (*p* < .05).

**Table 3.** Results of the Linear Mixed Model Examining Variables Across Days of the Race in Male Cyclists.

| Variable | df | *F* | *p* value |
| --- | --- | --- | --- |
| Sleep onset (hh:mm) | 1, 25 | 2.49 | <.001 |
| Sleep offset (hh:mm) | 1, 25 | 2.02 | <.005 |
| Time in bed (h) | 1, 25 | 2.83 | <.001 |
| Total sleep time (h) | 1, 25 | 2.05 | .004 |
| Sleep efficiency (%) | 1, 25 | 0.36 | .998 |
| Light sleep (%) | 1, 25 | 0.39 | .996 |
| Slow wave sleep (%) | 1, 25 | 0.52 | .972 |
| REM sleep (%) | 1, 25 | 0.56 | .954 |
| Daily strain (au) | 1, 25 | 12.88 | <.001 |
| rMSSD (ms) | 1, 25 | 0.95 | .537 |
| Resting heart rate (beats·min<sup>-1</sup>) | 1, 25 | 0.57 | .946 |

Linear mixed models were used to compare variables across days of racing. The fixed effect in the models (i.e., 'day of race') included baseline (i.e., average value for each participant calculated from Monday to Sunday at least one week prior to the first day of the race), day of race (21 race days, 1 transfer day and 2 rest days) and post-race (i.e., average value for each participant calculated from post-race night 2 to post-night 9). au = arbitrary units, REM = rapid eye movement, rMSSD = root mean square of successive differences.

#### 3.2.3 Week of Race (Fig. 2–4)

The ‘week of race’ analyses included baseline (i.e., average value for each participant calculated from Monday to Sunday at least one week prior to the first day of the race), week 1 (stages 1 to 9 but excluding the transfer day), week 2 (stages 10 to 15 but excluding the first rest day) and week 3 (stages 16 to 20 but excluding the second rest day). The variables obtained on the night of the final stage (stage 21) were excluded from the analyses. There was no effect of ‘week of race’ on any of the sleep or heart rate variables except for daily strain (Table 4). Compared with baseline, daily strain was higher during week 1 (+5.6 au; *p* <.001), week 2 (+6.6 au; *p* <.001), and week 3 (+4.5 au; *p* = .003).

**Table 4.** Results of the Linear Mixed Model Examining Variables Across Weeks of the Race in Male Cyclists.

| Variable | df | *F* | *p* value |
| --- | --- | --- | --- |
| Sleep onset (hh:mm) | 1, 3 | 0.50 | .684 |
| Sleep offset (hh:mm) | 1, 3 | 1.46 | .228 |
| Time in bed (h) | 1, 3 | 0.28 | .841 |
| Total sleep time (h) | 1, 3 | 0.85 | .470 |
| Sleep efficiency (%) | 1, 3 | 0.80 | .496 |
| Light sleep (%) | 1, 3 | 1.28 | .285 |
| Slow wave sleep (%) | 1, 3 | 0.34 | .800 |
| REM sleep (%) | 1, 3 | 2.33 | .077 |
| Daily strain (au) | 1, 3 | 10.44 | <.001 |
| rMSSD (ms) | 1, 3 | 1.14 | .337 |
| Resting heart rate (beats·min<sup>-1</sup>) | 1, 3 | 1.13 | .339 |

Linear mixed models were used to compare variables across the weeks of the race. The fixed effect in the models (i.e., ‘week of race’) included baseline (i.e., average value for each participant calculated from Monday to Sunday at least one week prior to the first day of the race), week 1 (stages 1 to 9 but excluding the transfer day), week 2 (stages 10 to 15 but excluding the first rest day) and week 3 (stages 16 to 20 but excluding the second rest day). au = arbitrary units, REM = rapid eye movement, rMSSD = root mean square of successive differences.

#### 3.2.4 Stage Classification (Fig. 5)

On average, participants covered 193.3 ± 2.7 km during flat stages, 179.5 ± 6.2 km during hilly stages, 161.4 ± 6.4 km during mountain stages, and 25.0 ± 7.4 km during time trials. On average, it took participants 4.1 ± 0.2 hours to complete flat stages, 4.2 ± 0.2 hours to complete hilly stages, 4.6 ± 0.2 hours to complete mountain stages, and 0.5 ± 0.2 hours to complete the time trials. There was a main effect of ‘stage classification’ on heart rate variability and daily strain (Table 5). Heart rate variability was lower (-23.9 ms; *p* = .010) following mountain stages than following rest days. Compared with rest days, daily strain was higher for flat stages (+5.8 au; *p* <.001), hilly stages (+7.1 au; *p* <.001), and mountain stages (+7.6 au; *p* <.001). Compared with individual time trials, daily strain was higher for flat stages (+7.2 au; *p* <.001), hilly stages (+8.6 au; *p* <.001), and mountain stages (+9.0 au; *p* <.001). Compared with flat stages, daily strain was higher for mountain stages (+1.7 au; *p* = .017). There was a main effect of ‘stage classification’ on time in bed (Table 5). Time in bed was shortest following the individual time trials compared to the other stage classifications, but none of the pairwise comparisons were significant.

**Fig. 5.**
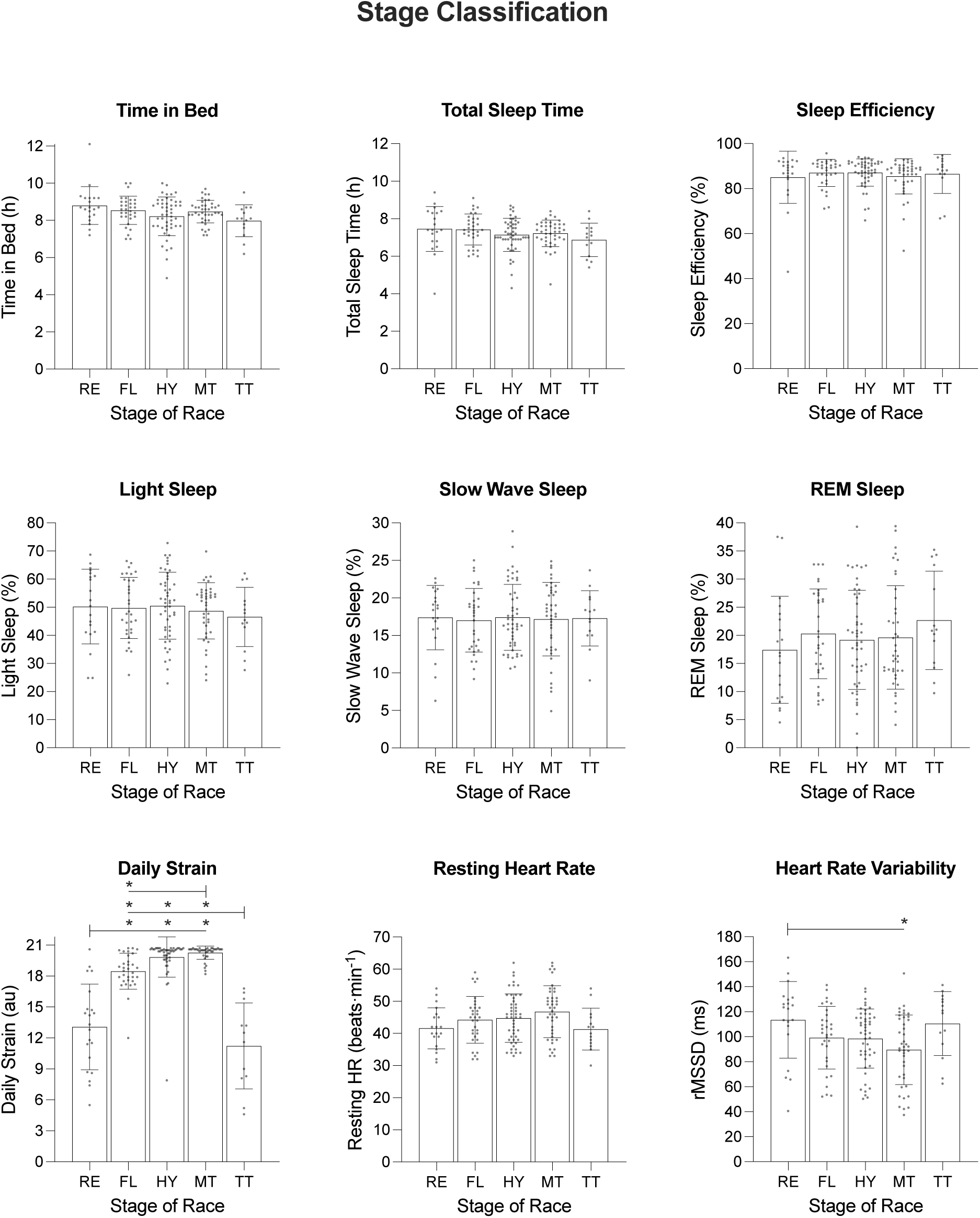
Sleep, heart rate, and daily strain of male cyclists during the Tour de France plotted as a function of stage classification. ‘RE’ indicates rest days; ‘FL’ indicates flat stages; ‘HY’ indicates hilly stages; ‘MT’ indicates mountain stages; and ‘TT’ indicates time trials. Data are presented as mean (bars) and 95% confidence intervals (error bars). Closed circles represent individual cyclists. The outcomes of the post hoc comparisons between stage classification are presented in each panel. * indicates a significant difference between stage classifications (*p* < .05).

**Table 5.**
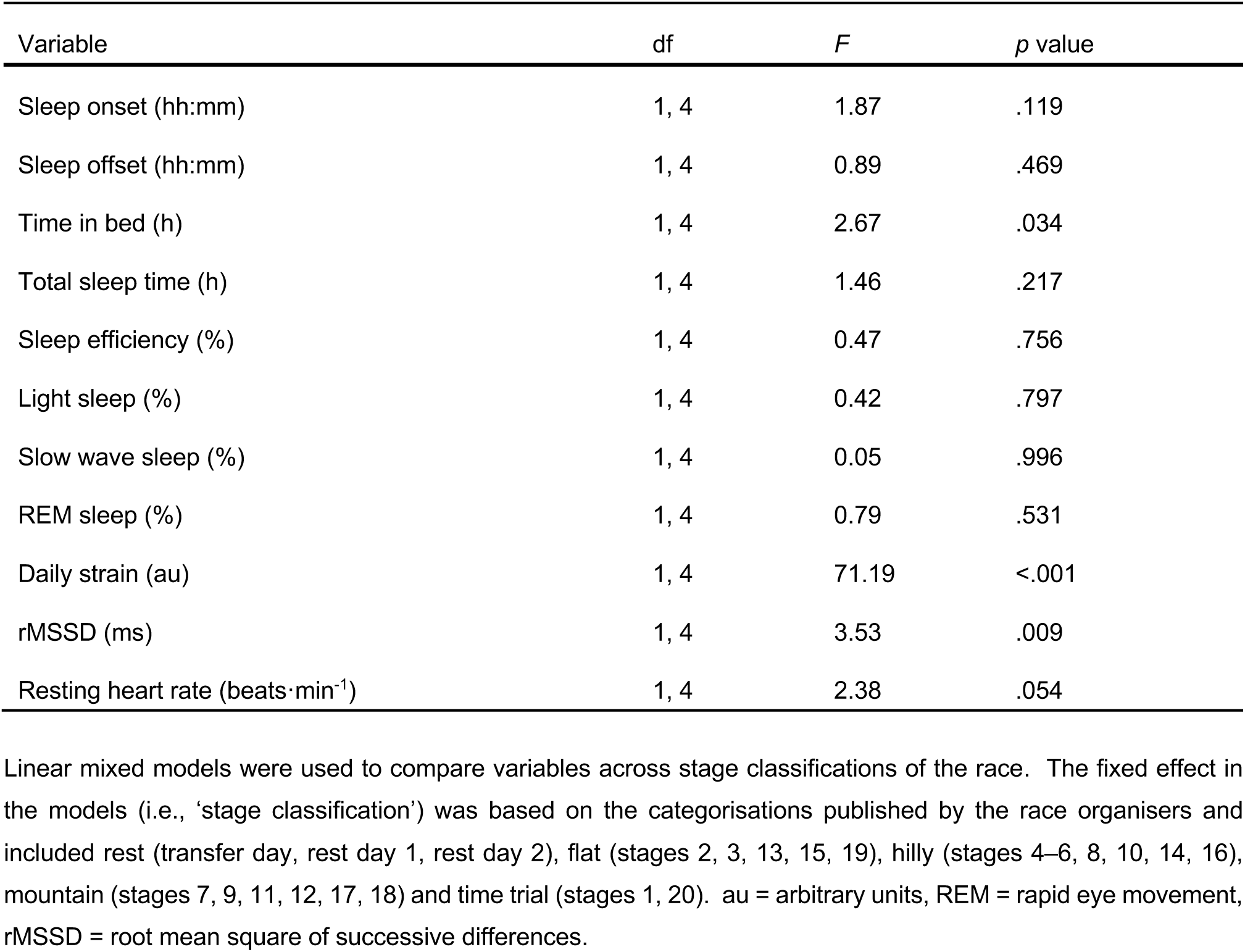
Results of the Linear Mixed Model Examining Variables Across Stage Classifications in Male Cyclists.

### 3.4 Female Cyclists

#### 3.3.1 Baseline

For each participant, data were collected at least one week prior to the start of the first stage of the race and included five weekdays (Monday-Friday) and two weekend days (Saturday, Sunday). On average, the participants fell asleep at 21:18 h ± 26.4 min, woke at 05:58 h ± 20.4 min, spent 8.7 ± 0.4 h in bed, and obtained 7.7 ± 0.3 h of sleep with an efficiency of 88.8 ± 2.6 %. The percentage of time in bed spent in each stage of sleep was 41.9 ± 3.6 % for light sleep, 20.2 ± 1.8 % for slow wave sleep and 26.7 ± 4.0 % for REM sleep. In comparison, for male/female non-athletes of a similar age (25 years), the percentage of time in bed spent in each stage of sleep is 54.8 % for light sleep, 16.3 % for slow wave sleep and 20.7 % for REM sleep [19]. The participants’ average resting heart rate was 45.8 ± 4.9 beats·min^-1^ and average heart rate variability was 119.8 ± 26.4 ms. In comparison, male/female non-athletes of a similar age (20–29 years) have resting heart rate of 54.2 beats·min^-1^ and heart rate variability of 82.9 ms [20].

#### 3.3.2 Day of Race (Fig. 6–9)

On average during the race, the participants fell asleep at 21:37 h ± 18.6 min, woke at 06:03 h ± 10.8 min, spent 8.4 ± 0.3 h in bed, and obtained 7.5 ± 0.3 h of sleep with an efficiency of 89.6 ± 1.2 %. The percentage of time in bed spent in each stage of sleep was 47.0 ± 2.6 % for light sleep, 19.3 ± 1.3 % for slow wave sleep and 23.3 ± 2.2 % for REM sleep. The participants’ average resting heart rate was 50.2 ± 2.0 beats·min^-1^ and average heart rate variability was 114.3 ± 11.2 ms. There was an effect of ‘day of race’ on sleep onset, time in bed, total sleep time, percentage of light sleep, percentage of REM sleep, and daily strain (Table 6). Time in bed and total sleep time gradually decreased over the course of the race and the percentage of light sleep increased over the course of the race. Specifically, participants fell asleep later (+3.1 h; *p* <.001), spent less time in bed (-2.5h; *p* <.001), and obtained less sleep (-2.2 h; *p* <.001) on the night after completing stage 8 compared with baseline; and the percentage of light sleep obtained was higher following stages 7 (+13.0%; *p* = .043) and 8 (+14.1%; *p* = .0030) compared with baseline. The percentage of REM sleep declined over the course of the race, but none of the pairwise comparisons were significant (reference category = baseline). Compared with baseline, daily strain was higher following stage 1 (*p* < .001), stage 2 (*p* < .001), stage 3 (*p* < .001), stage 4 (*p* < .001), stage 5 (*p* < .001), stage 6 (*p* < .001), stage 7 (*p* < .001) and stage 8 (*p* = .009).

**Fig. 6.**
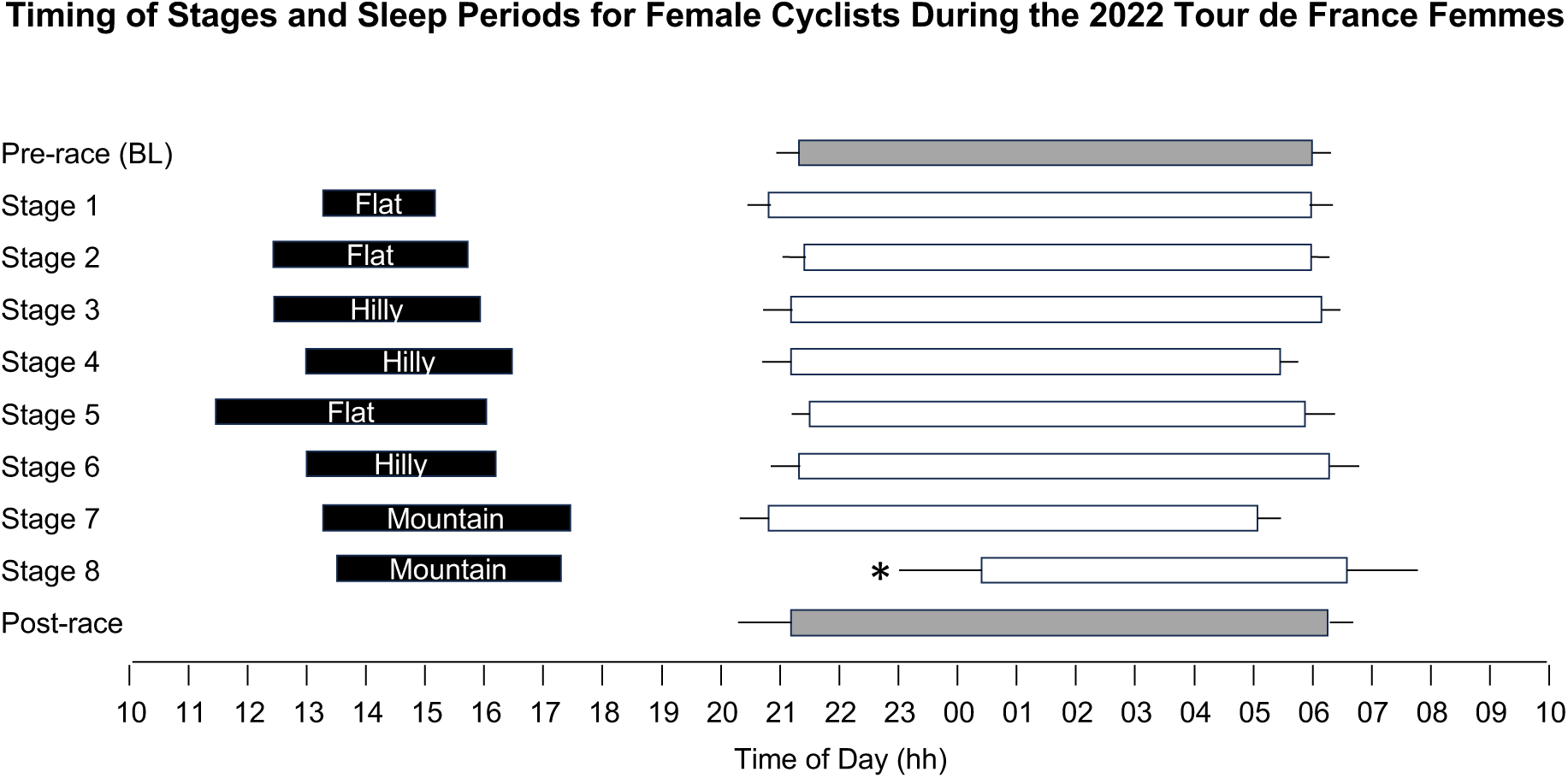
Timing of stages and sleep periods for female cyclists during the 2022 Tour de France Femmes. Each line represents a 24-hour day from 10:00 AM to 10:00 AM. Black bars indicate the start/end times and duration of stages, based on the cyclists who participated in the study. The start/end times and duration of sleep periods for participating cyclists are represented by white bars for sleep after race stages and dark grey bars for sleep in the 14 days before and after competition. Data are mean and 95% confidence intervals. * indicates a significant difference from baseline (*p* < .05).

**Fig. 7.**
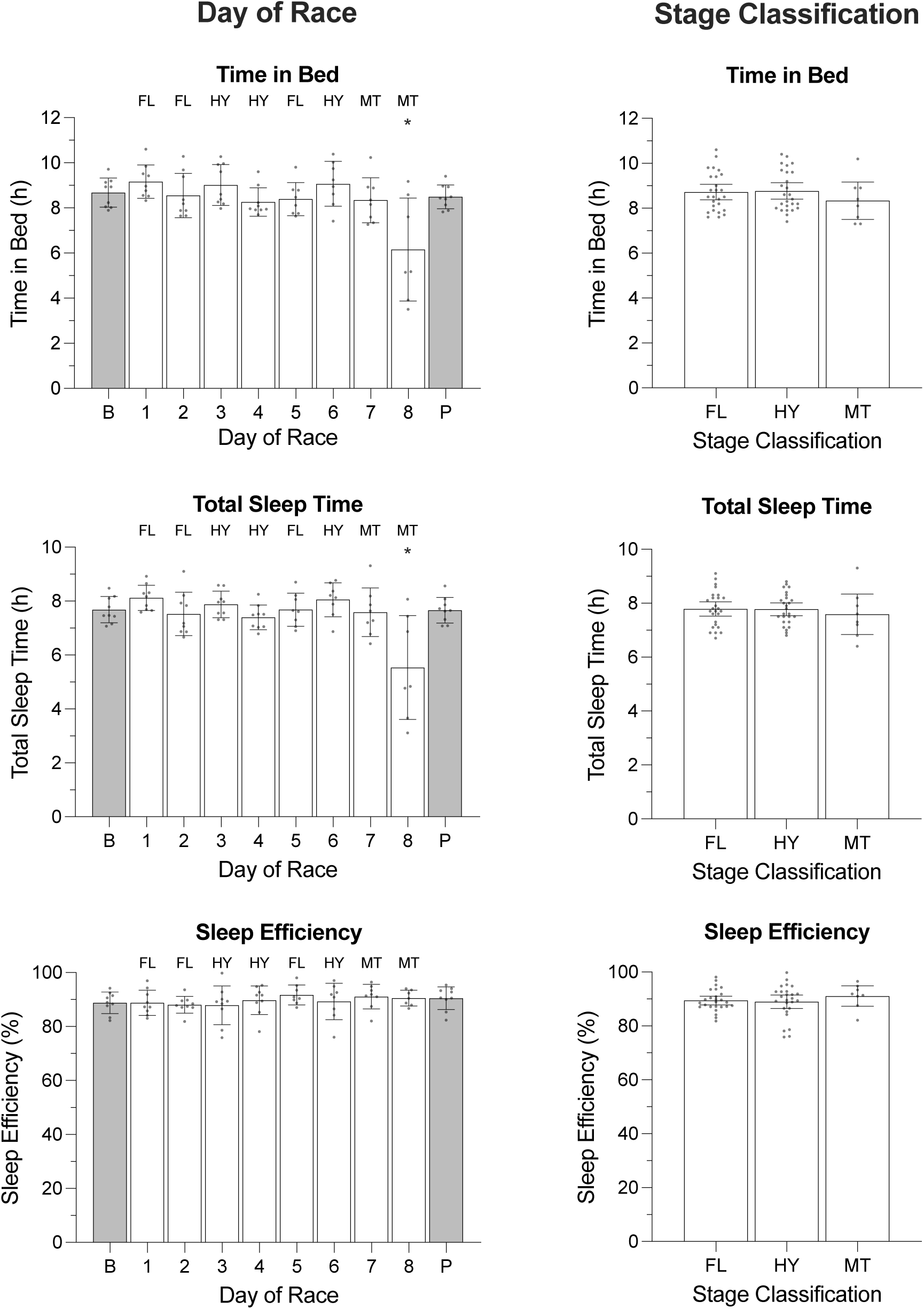
Time in bed, total sleep time, and sleep efficiency of female cyclists during the Tour de France Femmes plotted as a function of day of race (left panel) and stage classification (right panel). ‘B’ represents the baseline average calculated at least one week prior to the first day of the race; ‘P’ indicates the post-race average calculated from post-race night 2 to post-night 9; ‘FL’ indicates flat stages; ‘HY’ indicates hilly stages; ‘MT’ indicates flat stages; ‘H’ indicates hilly stages; ‘M’ indicates mountain stages. Data are presented as mean (bars) and 95% confidence intervals (error bars). Closed circles represent individual cyclists. In the left panel, * indicates a significant difference from baseline (*p* < .05).

**Fig. 8.**
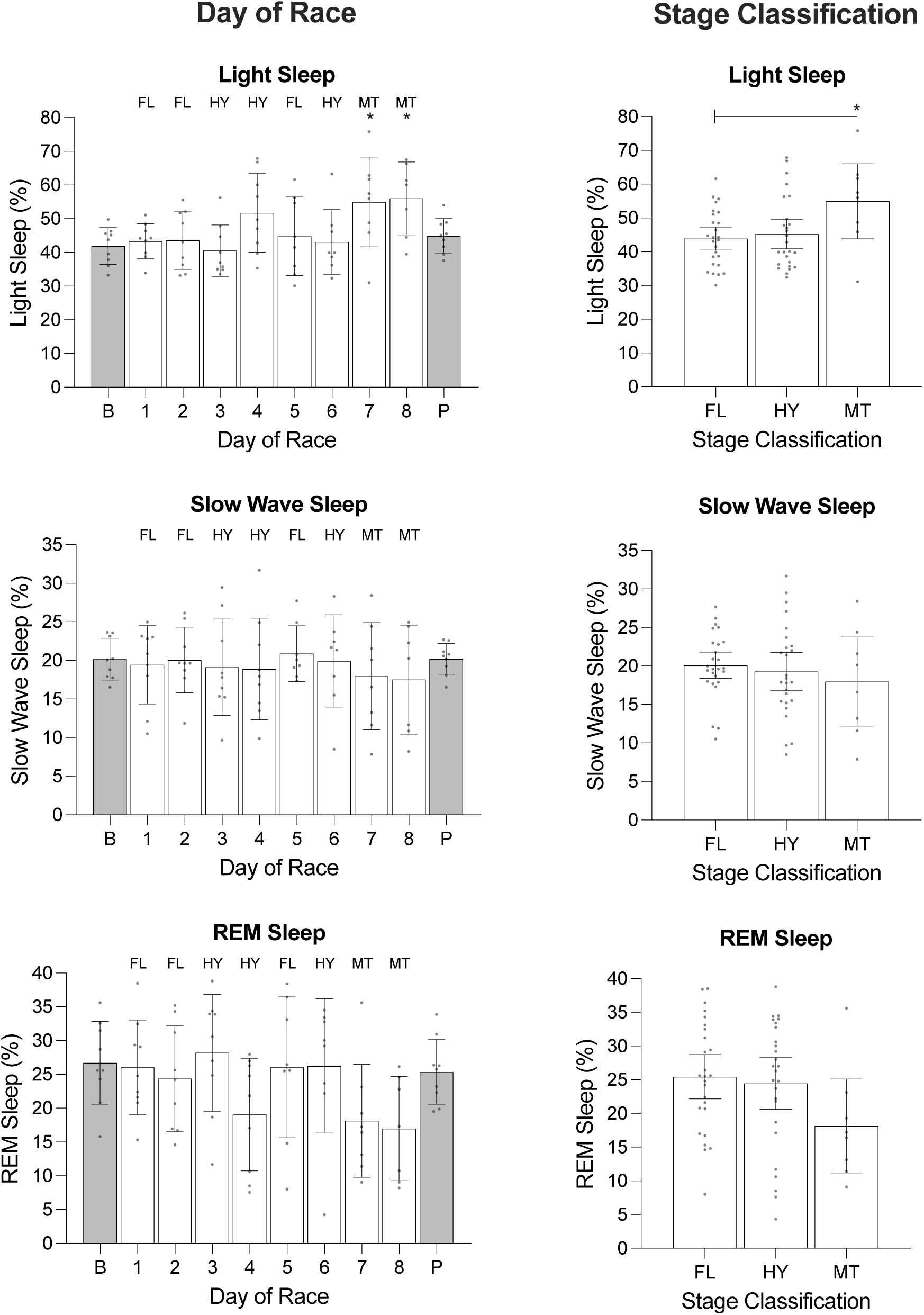
The percentage of light sleep, slow wave sleep and REM sleep obtained by female cyclists during the Tour de France Femmes plotted as a function of day of race (left panel) and stage classification (right panel). ‘B’ represents the baseline average calculated at least one week prior to the first day of the race; ‘P’ indicates the post-race average calculated from post-race night 2 to post-night 9; ‘FL’ indicates flat stages; ‘HY’ indicates hilly stages; ‘MT’ indicates mountain stages. Data are presented as mean (bars) and 95% confidence intervals (error bars). Closed circles represent individual cyclists. In the left panel, * indicates a significant difference from baseline (*p* < .05). In the right panel, * indicates a significant difference between stage classifications (*p* < .05).

**Fig. 9.**
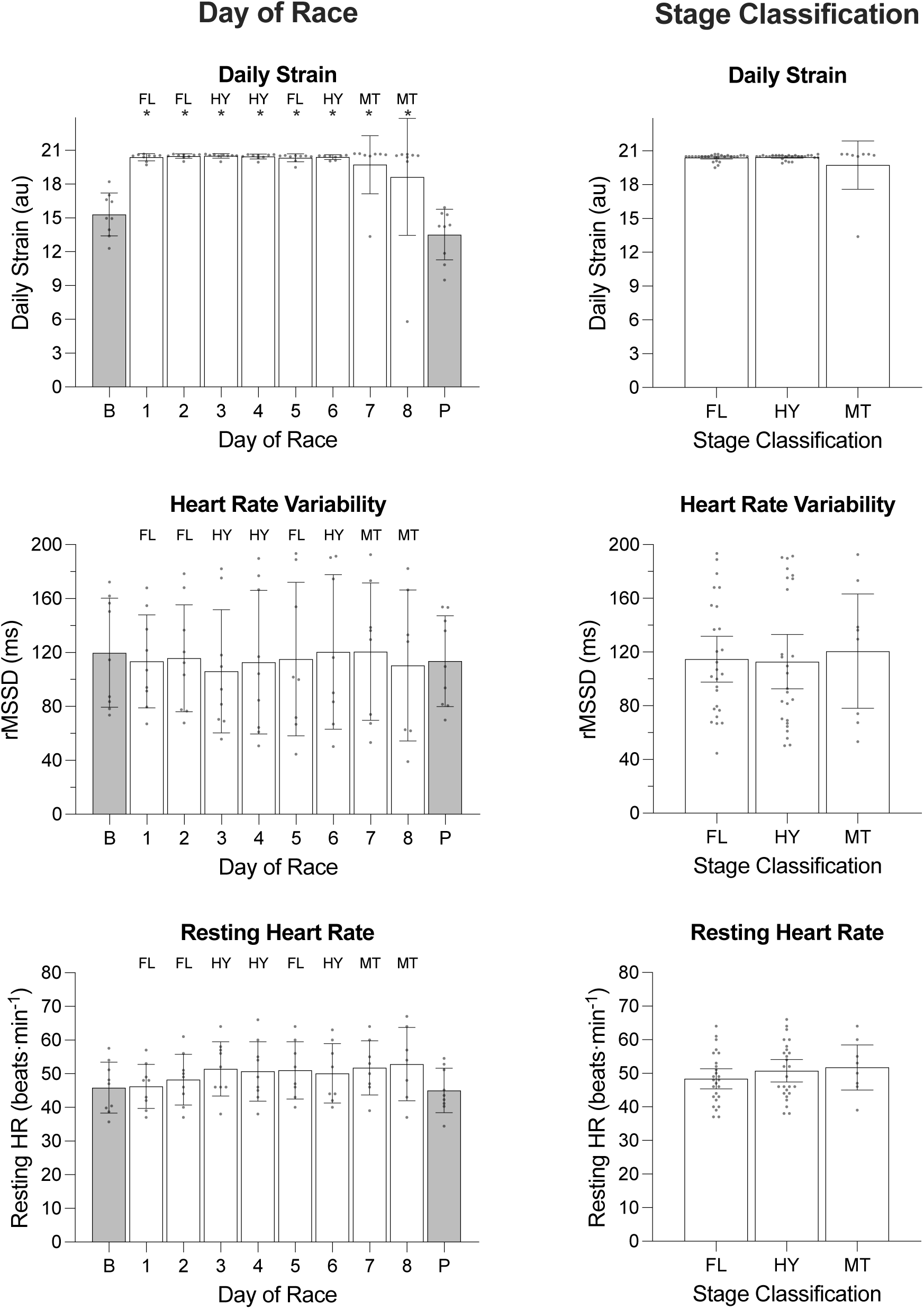
Daily strain, heart rate variability and resting heart rate of female cyclists during the Tour de France Femmes plotted as a function of day of race (left panel) and stage classification (right panel). ‘B’ represents the baseline average calculated at least one week prior to the first day of the race; ‘P’ indicates the post-race average calculated from post-race night 2 to post-night 9; ‘FL’ indicates flat stages; ‘HY’ indicates hilly stages; ‘MT’ indicates mountain stages. Data are presented as mean (bars) and 95% confidence intervals (error bars). Closed circles represent individual cyclists. In the left panel, * indicates a significant difference from baseline (*p* < .05).

**Table 6.** Results of the Linear Mixed Model Examining Variables Across Days of the Race in Female Cyclists.

| Variable | df | *F* | *p* value |
| --- | --- | --- | --- |
| Sleep onset (hh:mm) | 1, 9 | 10.01 | <.001 |
| Sleep offset (hh:mm) | 1, 9 | 1.47 | .176 |
| Time in bed (h) | 1, 9 | 5.26 | <.001 |
| Total sleep time (h) | 1, 9 | 6.19 | <.001 |
| Sleep efficiency (%) | 1, 9 | 0.60 | .795 |
| Light sleep (%) | 1, 9 | 2.96 | .005 |
| Slow wave sleep (%) | 1, 9 | 0.32 | .968 |
| REM sleep (%) | 1, 9 | 2.07 | .043 |
| Daily strain (au) | 1, 9 | 14.16 | <.001 |
| rMSSD (ms) | 1, 9 | 0.08 | 1.00 |
| Resting heart rate (beats·min<sup>-1</sup>) | 1, 9 | 0.98 | .461 |

Linear mixed models were used to compare variables across days of racing. The fixed effect in the models (i.e., 'day of race') included baseline (i.e., average value for each participant calculated from Monday to Sunday at least one week prior to the first day of the race), day of race (8 race days) and post-race (i.e., average value for each participant calculated from post-race night 2 to post-night 9). au = arbitrary units, REM = rapid eye movement, rMSSD = root mean square of successive differences.

#### 3.3.3 Stage Classification (Fig. 7–9)

On average, participants covered 131.2 ± 14.8 km during flat stages, 129.7 ± 1.2 km during hilly stages, and 125.1 ± 1.0 km during the mountain stages. On average, it took participants 3.3 ± 0.4 hours to complete flat stages, 3.4 ± 0.1 hours to complete hilly stages, and 4.0 ± 0.1 hours to complete mountain stages. There was no effect of ‘stage classification’ on any of the sleep or heart rate variables except for the percentage of light sleep obtained (Table 7). The percentage of light sleep was higher (+11.4 %) following the mountain stage compared with flat stages (*p* = .036).

**Table 7.** Results of the Linear Mixed Model Examining Variables Across Stage Classifications in Female Cyclists.

| Variable | df | *F* | *p* value |
| --- | --- | --- | --- |
| Sleep onset (hh:mm) | 1, 2 | 2.04 | .140 |
| Sleep offset (hh:mm) | 1, 2 | 0.14 | .873 |
| Time in bed (h) | 1, 2 | 0.74 | .483 |
| Total sleep time (h) | 1, 2 | 0.30 | .739 |
| Sleep efficiency (%) | 1, 2 | 0.51 | .601 |
| Light sleep (%) | 1, 2 | 3.76 | .029 |
| Slow wave sleep (%) | 1, 2 | 0.48 | .619 |
| REM sleep (%) | 1, 2 | 2.17 | .124 |
| Daily strain (au) | 1, 2 | 1.96 | .150 |
| rMSSD (ms) | 1, 2 | 0.08 | .919 |
| Resting heart rate (beats·min<sup>-1</sup>) | 1, 2 | 0.86 | .430 |

Linear mixed models were used to compare variables across stage classifications of the race. The fixed effect in the models (i.e., 'stage classification') was based on the categorisations published by the race organisers and included flat (stages 1, 2, 5), hilly (stages 3, 4, 6), and mountain (stages 7, 8). au = arbitrary units, REM = rapid eye movement, rMSSD = root mean square of successive differences.

## 4 Discussion

In the present study, sleep and autonomic activity were monitored in professional male and female cyclists during the Tour de France and Tour de France Femmes, respectively. The aims of the study were to (i) quantify the impact of days of the race on sleep and autonomic activity; (ii) examine whether the characteristics of stages of the race differentially affect sleep and autonomic activity; and (iii) determine whether there is a cumulative effect of racing (i.e., over weeks) on sleep and autonomic activity. The major findings of the study are that (i) the amount of sleep obtained by male and female cyclists did not change over the course of the race nor did heart rate variability; (ii) sleep quality was poorer after a mountain stage compared to flat stages in female cyclists; (iii) heart rate variability was lower after mountain stages compared to flat stages in male cyclists; and (iv) there was no cumulative effect of racing on sleep or heart rate variability in male cyclists.

There is some evidence to indicate that sleep is reduced on nights before and after competition [21–24] and that endurance exercise can also disrupt subsequent sleep [9]. In this regard, the Tour de France and the Tour de France Femmes are unique events because cyclists complete prolonged bouts of endurance exercise over successive days of competition. Contrary to expectations, total sleep time in the present study was not influenced by consecutive days of racing and remained relatively stable across most nights of the race. In addition, average total sleep time for both male (7.2 h) and female (7.5 h) cyclists during their respective races was substantially higher than that reported for road cyclists during a normal phase of training (6.3 h) [10] and higher than that reported for road cyclists during one-week multi-stage races [11, 12] and a ‘simulated’ three-week Grand Tour [13]. There are several possible explanations for the good sleep observed in the present study. Given the importance and prestige of both the Tour de France and Tour de France Femmes, cyclists may have prioritised sleep over other activities to ensure optimal recovery and performance [25]. In addition, the schedule of racing may have been favourable in terms of cyclists’ opportunities for sleep. For example, the average start time of stages of the race was 13:00 h for both male and female cyclists – the earliest start time of a stage during the Tour de France was 12:10 h and the earliest start time of a stage during the Tour de France Femmes was 11:45 h. Later start times are associated with increased night-time sleep duration during training [26] and it seems reasonable to extrapolate that later start times during competition may also be associated with good night-time sleep (i.e., >7 h).

Compared to the male cyclists, female cyclists in the present study exhibited poorer sleep quality over the course of their respective race. Specifically, the percentage of light sleep was higher (+13.6 %), and the percentage of REM sleep was lower (-9.2 %), following the two mountain stages (stages 7 and 8) compared to baseline. Similar changes in light sleep and REM sleep have been observed following a single bout of ultra-endurance exercise in male athletes. For example, when compared to a day when no exercise was performed, light sleep increased (+27 min) and REM sleep decreased (-45 min) on the night following an ultra-triathlon [9]; and REM sleep was reduced (-6%) in well-trained cyclists following a single stage race (120-150 km) compared to a non-training/non-racing recovery period [27]. Prolonged endurance exercise can elevate core body temperature [27, 28], elicit muscle soreness [29, 30], and increase sympathetic activity [27] – all of which have the potential to disrupt sleep [9]. The probability that an increase in core body temperature is responsible for the changes in sleep observed in the present study may be small given that sleep opportunities began on average ∼5.5 h after stage completion, over which time core body temperature should progressively decrease [27, 31]. An increase in muscle soreness or an increase in sympathetic activity may provide more likely explanations for an increase in light sleep and a decrease in REM sleep. For example, pain increases the number of arousals during sleep and the sleep arousal threshold to pain is lowest during light sleep compared to slow wave sleep or REM sleep [32]; and catecholamine excretion during the night increases following prolonged endurance exercise [27, 33], which may suppress REM sleep [26, 32]. In future, it may be useful to measure muscle soreness or catecholamine excretion following stages to determine the extent to which these factors disrupt sleep.

Indices of heart rate and heart rate variability are tools that can be used to quantify the acute stress-recovery responses of the autonomic nervous system following a bout of exercise [34, 35]. In the present study, heart rate variability was lower after mountain stages than after rest days in male cyclists. A similar negative relationship between heart rate variability and physiological load has been observed in male cyclists competing in the Vuelta a España [7] and in female cyclists riding the route of the 2017 Tour de France [14]. In each of these studies, heart rate variability was lowest following either the most demanding stages (i.e., mountain) [14] or after periods of racing in which physiological load was very high [7]. Despite an acute reduction in heart rate variability following mountain stages in male cyclists, there was no cumulative effect of racing on heart rate variability in male or female cyclists. For male cyclists, the presence of rest days and the distribution of flat stages between hilly or mountain stages during the race could provide sufficient time over which to recover and prevent further reductions in heart rate variability. For female cyclists, shorter race duration and placement of mountain stages at the end of the race could explain why heart rate variability remained stable throughout the race.

In the present study, the primary analyses focussed on the impact of the race schedule (i.e., consecutive days of racing over flat, hilly, mountain stages) on subsequent recovery (i.e., sleep and heart rate variability). Team strategy is another major factor that could affect the physiological and psychological demands placed on riders during the race and their subsequent recovery following each stage. Teams competing in the Tour de France or Tour de France Femmes consist of eight and six riders, respectively – a team leader, other riders (i.e., domestiques), and stage specialists (i.e., sprinters, climbers). For each stage, riders may perform different roles depending on the performance of the team and the strategy of the team. For example, riders may be required to (i) set the pace at the front of the peloton to protect and conserve the energy of their team leader; (ii) defend their team leader against attacks from other riders; (iii) assist their team leader in initiating attacks against other riders to gain time or points; or (iv) form a breakaway to challenge for a stage win or support a teammate aiming for a stage win. These roles are likely to influence how well cyclists recover during the race. For example, heart rate variability was substantially lower for a domestique after the first 15 stages in the Tour of Spain compared to that of the team leader [7]. In the present study, there was insufficient data to examine the potential impact of individual and team strategies on subsequent recovery following each stage of the race. In future, it may be possible to accumulate data from multiple teams over multiple races to determine whether team strategy or team performance during a race affects subsequent recovery in terms of sleep and heart rate variability.

There are some limitations that should be considered when interpreting the results of the present study. First, it is impractical to use the gold-standard equipment for assessing sleep, i.e., polysomnography, with professional athletes during peak events such as the Tour de France and Tour de France Femmes, so alternative equipment must be employed. In this case, a wearable fitness tracker, i.e., WHOOP 4.0 was used to assess night-time sleep and autonomic activity. An earlier version of the device, i.e., WHOOP 3.0, has excellent agreement with gold-standard measures of heart rate and heart rate variability and moderate agreement with gold-standard measures of sleep [15]. As with other fitness trackers that assess sleep, WHOOP is very good at detecting sleep in general, but its capacity to identify wake that occurs within a sleep period, and its capacity to identify particular stages of sleep, could be further improved [15]. This should be considered when interpreting the results regarding sleep reported in the present study. Despite this caution regarding interpretation, this study demonstrates that it is now feasible to assess recovery metrics in professional athletes during multiple-day endurance events using validated fitness trackers. Second, compliance was very high on almost all nights before and after the race, but it was poorest on the night of the final stage of the race. For this reason, data on the night of the final stage was excluded from most analyses. For the male cyclists, this resulted in data being excluded from the final flat stage and for the female cyclists, this resulted in data being excluded from the final mountain stage. It is possible that exclusion of these data from some of the analyses may have altered the results. However, sleep/wake behaviours on the final night of racing may be influenced by factors other than the race (e.g., celebrations, media commitments, social demands, etc) and as such, may not accurately reflect recovery. Finally, napping is an effective strategy when athletes’ night-time sleep is restricted [36], and some athletes do nap during competition [37, 38], but naps were not included in the analyses in the present study.

## Data Availability

All data produced in the present study are available upon reasonable request to the authors.

## Declarations

### Acknowledgements

The authors are grateful to the cyclists and staff from the participating teams for the time and effort taken during data collection and for making the data available for publication.

### Ethics approval

The study was approved by CQUniversity’s Human Research Ethics Committee.

